# A Nationwide Survey of UK cardiac surgeons’ view on clinical decision making during the COVID-19 pandemic

**DOI:** 10.1101/2020.04.24.20078105

**Authors:** Umberto Benedetto, Andrew Goodwin, Simon Kendall, Rakesh Uppal, Enoch Akowuah

**Author notes:** Corresponding author Umberto Benedetto, Bristol Royal Infirmary, Upper Maudlin Street BS2 8HW. **No conflict to disclose**.

## Abstract

**Background:** No firm recommendations are currently available to guide decision making for patients requiring cardiac surgery during the COVID-19 pandemic. Systematic appraisal of national expert consensus can be used to generate interim recommendations until data from clinical observations will become available. Hence, we aimed to collect and quantitatively appraise nationwide UK senior surgeons’ opinion on clinical decision making for patients requiring cardiac surgery during the COVID-19 pandemic.

**Methods:** We mailed a web-based questionnaire to all consultant cardiac surgeons through the Society for Cardiothoracic Surgery in Great Britain and Ireland (SCTS) mailing list on the 17^th^ April 2020 and we pre-determined to close the survey on the 21^st^ April 2020. This survey was primarily designed to gather information on UK surgeons’ opinion using 12 items. Strong consensus was predefined as an opinion shared by at least 60% of responding consultants.

**Results:** A total of 86 consultant surgeons undertook the survey. All UK cardiac units were represented by at least one consultant. Strong consensus was achieved for the following key questions:1) before hospital admission every patient should receive nasopharyngeal swab, PCR and chest CT; 2) the use of full PPE should to be adopted in every case by the theatre team regardless patient’s COVID-19 status; 3) the risk of COVID-19 exposure for patients undergoing heart surgery should be considered moderate to high and likely to increase mortality if it occurs; 4) cardiac procedure should be decided based on ad-hoc multidisciplinary team discussion for every patient. The majority believed that both aortic and mitral surgery should be considered in selected cases. The role of CABG surgery during the pandemic was more controversial.

**Conclusions:** In the current unprecedented scenario, the present survey provides information for generating interim recommendations until data from clinical observations will become available.

**Perspective statement:** Systematic appraisal of national expert consensus can be used to generate interim recommendations for patients undergoing cardiac surgery during COVID-19 pandemic until data from clinical observations will become available.

**Central message:** No firm recommendations are currently available to guide decision making for patients requiring cardiac surgery during the pandemic. This can translate into significant variability in clinical practice and patients’ outcomes across cardiac units. Systematic appraisal of national expert consensus can represent a rapid and efficient instrument to provide support to heath policy makers and other stakeholders in generating interim recommendations until data from clinical observations will become available.

## Introduction

The Coronavirus disease 2019 (COVID-19) pandemic has had an unprecedented impact on healthcare globally, including on the delivery of cardiac surgical care [1-2]. Cardiac surgery is the single largest user of intensive care unit beds [1-2] and resource re-allocation to treat COVID-19 patients has halted the provision of routine cardiac surgery services worldwide and in the United Kingdom. Despite the disruption, urgent and emergency cardiac procedures are still required by the public during the pandemic. However, there remain several areas of uncertainty, including the risks incurred by patients with heart conditions, who may suffer fatal events if cardiac surgery is delayed by several weeks, and the impact of acquiring COVID-19 during the perioperative period. Anecdotal evidence suggest post-operative COVID-19 infection may be fatal.

No firm recommendations are currently available to guide decision making for patients requiring cardiac surgery during the pandemic. This can translate into significant variability in clinical practice and patients’ outcomes across cardiac units. In these circumstances, expert consensus can provide interim guidance for healthcare policy makers, for clinicians’ daily practice and for patients [3]. We aimed to collect and quantitatively appraise nationwide UK senior surgeons’ opinion on clinical decision making for patients requiring cardiac surgery during the COVID-19 pandemic.

### Participants and methods

We mailed a web-based questionnaire to all consultant cardiac surgeons through the Society for Cardiothoracic Surgery in Great Britain and Ireland (SCTS) mailing list on the 17^th^ April 2020. In view of the rapidly evolving circumstances and the need for timely outcome presentation, we pre-determined to close the survey on the 21^st^ April 2020. This survey was primarily designed to gather information on UK surgeons’ opinion on which patients should be considered for cardiac surgery under the current COVID-19 pandemic using 12 items. As at the time of the survey, there was significant variability on clinical activities across centres, the first part of the questionnaire gathered information on local factors (local resource relocation to treat COVID-19) that may have influenced surgeons’ view. Strong consensus was predefined as an opinion shared by at least 60% of responding consultants [3].

## Results

A total of 86 consultant surgeons undertook the survey. All UK cardiac units were represented by at least one consultant. Figure 1 shows the distribution of responding consultants across different regions and the proportion of consultant stratified by local resource relocation. Geographical regions with the highest number of responding consultant were London area, North West and Northern regions. Most consultants were from units where resources were only partially redirected to treat COVID-19 (n=63, 73%) followed by consultants working in units entirely relocated (n=17, 18%) and only nine consultants were from in units where resources were not redirected (10%). Table 1 shows the results of the survey in the overall sample and in groups stratified by working in units with resource relocation.

**Table 1.** Results of the survey among 86 consultant cardiac surgeons (at least one from each UK unit) in the overall sample and stratified by resource relocation to treat COVID-19. (strong consensus highlighted in yellow, majority in bold; COVID-19: Coronavirus disease 2019; PRC: polymerase chain reaction; CT computerised tomography; PPE Personal Protective Equipment; MDT: multidisciplinary team; STEMI: ST-elevation myocardial infarction; CABG: coronary artery bypass graft; AV: aortic valve; MV: mitral valve; TAVI: Transcatheter aortic valve implantation)

| Screening for COVID-19 before patient's admission for non-salvage cardiac surgery should consist of: | Total | Resource relocated |  |  |
| --- | --- | --- | --- | --- |
|  |  | No | Partially | Entirely |
| I do not know | 1.2% | 0.0% | 1.6% | 0.0% |
| nasopharyngeal swab and PCR for suspected cases only | 1.2% | 12.5% | 0.0% | 0.0% |
| **nasopharyngeal swab, PCR and chest CT for every patient** | **60.5%** | 62.5% | 65.1% | 40.0% |
| nasopharyngeal swab, PCR and chest CT for suspected case only | 5.8% | 0.0% | 6.3% | 6.7% |
| nasopharyngeal swab, PCR for every patient. | 31.4% | 25.0% | 27.0% | 53.3% |
| **During this pandemic, full PPE should be adopted by the theatre team:** |  |  |  |  |
| I don't know | 1.2% | 0.0% | 1.6% | 0.0% |
| **In every case regardless patient COVID-19 status** | **60.5%** | 62.5% | 54.0% | 86.7% |
| Only in a confirmed COVID-19 case or in all cases where COVID-19 screening was not performed | 17.4% | 12.5% | 22.2% | 0.0% |
| Only in a confirmed or suspect COVID-19 case | 20.9% | 25.0% | 22.2% | 13.3% |
| **During this pandemic, the risk of COVID-19 exposure for patients undergoing cardiac surgery is:** |  |  |  |  |
| I don't know | 3.5% | 0.0% | 3.2% | 6.7% |
| Low but likely to increase mortality if it occurs | 25.6% | 12.5% | 28.6% | 20.0% |
| **Moderate to high and likely to increase mortality if it occurs** | **69.8%** | 87.5% | 66.7% | 73.3% |
| Moderate to high but unlikely to increase mortality if it occurs | 1.2% | 0.0% | 1.6% | 0.0% |
| **During this pandemic, cardiac surgery operations should be performed:** |  |  |  |  |
| As usual following standard recommendations | 9.3% | 0.0% | 11.1% | 6.7% |
| At surgeons' discretions | 12.8% | 12.5% | 12.7% | 13.3% |
| I don't know | 1.2% | 0.0% | 1.6% | 0.0% |
| Only after ad-hoc MDT for every case | 64.0% | 50.0% | 65.1% | 66.7% |
| Surgery should never be performed unless strictly necessary (i.e. dissection) | 12.8% | 37.5% | 9.5% | 13.3% |
| **a patient confirmed or suspected COVID-19 positive presenting with acute type A dissection should be operated on:** |  |  |  |  |
| I don't know | 2.3% | 12.5% | 0.0% | 6.7% |
| Only if he/she has no symptoms of infection (i.e. no fever, normal blood cell count, normal chest CT) | 22.1% | 12.5% | 23.8% | 20.0% |
| Only if he/she has no symptoms of infection and has best chances of survival (i.e. age) | 53.5% | 50.0% | 60.3% | 26.7% |
| Should be considered for surgery only if he/she is unstable (i.e. cardiac tamponade) | 17.4% | 12.5% | 15.9% | 26.7% |
| Surgery should never be attempted | 4.7% | 12.5% | 0.0% | 20.0% |
| **During this pandemic elective surgery for non-COVID patients should be performed:** |  |  |  |  |
| All elective cases with priority (i.e. symptoms) to be considered for TAVI or PCI and surgery to be performed only if strictly necessary | 40.7% | 0.0% | 42.9% | 53.3% |
| As usual following standard recommendations | 2.3% | 0.0% | 3.2% | 0.0% |
| Only in cases with priority (i.e. symptoms) | 47.7% | 62.5% | 46.0% | 46.7% |
| Surgery should never be performed | 9.3% | 37.5% | 7.9% | 0.0% |
| **During this pandemic, surgery for non-COVID inpatients should be performed:** |  |  |  |  |
| All inpatients to be considered for TAVI or PCI and surgery to be performed only if strictly necessary | 40.7% | 12.5% | 39.7% | 60.0% |
| As usual following standard recommendations | 11.6% | 12.5% | 14.3% | 0.0% |
| Only in selected cases (age criteria, anatomy) | 45.3% | 50.0% | 46.0% | 40.0% |
| Surgery should never be performed | 2.3% | 25.0% | 0.0% | 0.0% |
| **During this pandemic, CABG surgery for non-COVID patients should be performed:** |  |  |  |  |
| As usual following standard recommendations | 4.7% | 12.5% | 4.8% | 0.0% |
| Neither CABG nor PCI should be performed unless strictly necessary (i.e. STEMI, unstable angina) | 22.1% | 12.5% | 22.2% | 26.7% |
| Only in selected cases (i.e. age criteria, left main disease) | 40.7% | 50.0% | 44.4% | 20.0% |
| PCI should always be the default strategy and CABG should be considered only in unstable patients when PCI is not feasible | 32.6% | 25.0% | 28.6% | 53.3% |
| **During this pandemic AV surgery for non-COVID patients should be performed:** |  |  |  |  |
| Following standard recommendations | 5.8% | 12.5% | 4.8% | 6.7% |
| Neither AV surgery nor TAVI should be performed unless strictly necessary (unstable or very symptomatic patients) | 30.2% | 25.0% | 31.7% | 26.7% |
| Only in selected cases (i.e. age criteria, bicuspid valve) | 51.2% | 62.5% | 54.0% | 33.3% |
| TAVI should always be the default strategy and AV surgery should be considered only in unstable patients when TAVI is not feasible | 12.8% | 0.0% | 9.5% | 33.3% |
| **During this pandemic MV surgery for non-COVID patients should be performed:** |  |  |  |  |
| Following standard recommendations | 3.5% | 12.5% | 3.2% | 0.0% |
| I don't know | 2.3% | 0.0% | 0.0% | 13.3% |
| MV surgery should never be performed unless strictly necessary (unstable or very symptomatic patients) | 41.9% | 37.5% | 38.1% | 60.0% |
| Only in selected cases (i.e. age criteria, very symptomatic) | 52.3% | 50.0% | 58.7% | 26.7% |
| **After this pandemic, which of the following sentence will be true?** |  |  |  |  |
| Cardiac surgery activities will be significantly reduced in favour of interventional procedures (i.e. TAVI, PCI) | 10.5% | 12.5% | 9.5% | 13.3% |
| Cardiac surgery activities will go back to normal | 65.1% | 62.5% | 65.1% | 66.7% |
| I don't know | 24.4% | 25.0% | 25.4% | 20.0% |
| **After this pandemic, future indications need be revised accounting for other factors (i.e. ICU beds utilization)** |  |  |  |  |
| I don't know | 8.1% | 12.5% | 7.9% | 6.7% |
| No | 68.6% | 75.0% | 66.7% | 73.3% |
| Yes | 23.3% | 12.5% | 25.4% | 20.0% |

**Figure 1.**
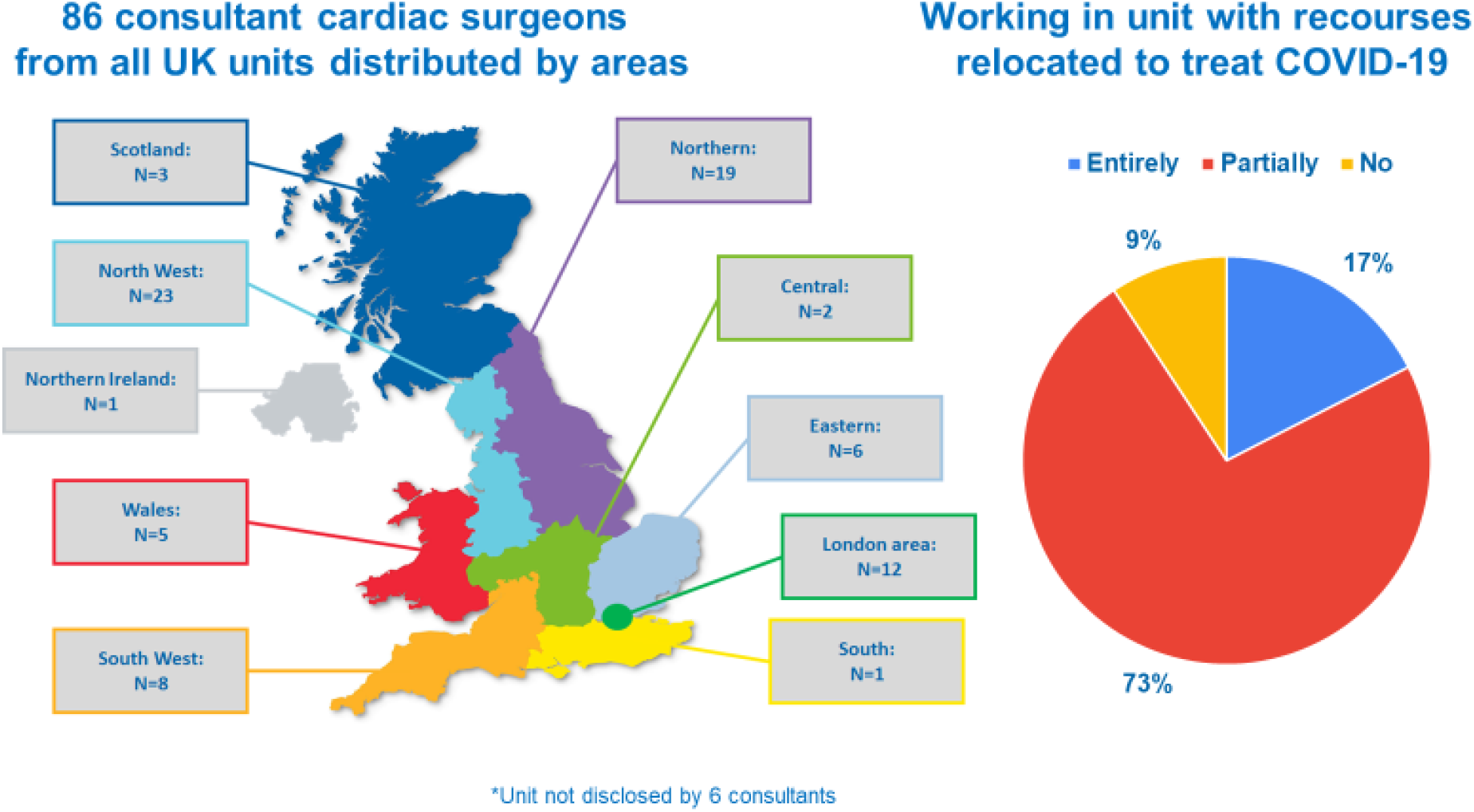
Distribution of responding consultants across macro-areas and proportion of responders working in units stratified by resources relocation to treat COVID-19

In the overall sample, strong consensus (≥60%) was achieved for the following key questions:1) before hospital admission every patient should receive nasopharyngeal swab, polymerase chain reaction (PCR) and chest computerised tomography (CT); 2) the use of full Personal Protective Equipment (PPE) should to be adopted in every case by the theatre team regardless patient’s COVID-19 status; 3) the risk of COVID-19 exposure for patients undergoing heart surgery should be considered moderate to high and likely to increase mortality if it occurs; 4) cardiac procedure should be decided based on ad-hoc multidisciplinary team (MDT) for every patient. Although there was no strong consensus on other key questions, the majority (>50%) agreed on that: 1) patients tested COVID-19 positive before salvage surgery (i.e. dissection), should be considered for surgery only if they have no symptoms of infection and have best chances of survival (taking into consideration factors such as age); 2) aortic and mitral valve surgery could similarly be considered only in selected cases. Interestingly, opinion about who should have coronary artery bypass grafting (CABG) surgery was much more varied. When the outcomes of the survey were stratified by resource relocation, surgeons from units where resources were not relocated (i.e. units which are carrying on as normal) showed a very strong agreement (>85%) that the risk of COVID-19 exposure for patients undergoing cardiac surgery is moderate to high and likely to increase mortality if it occurs. This group also showed the highest proportion of surgeons believing that cardiac surgery should never be performed in elective or urgent inpatients. Finally, there was a strong consensus that this pandemic will not have an impact on surgical activities when normal operating conditions will be re-established.

### Comments

We are realising that non COVID-19 infection related deaths may be extremely important component of the health impact of the COVID-19 pandemic worldwide due to healthcare resources relocation to acutely respond to the pandemic. However, there is little direct evidence to inform the management of patients requiring cardiac surgery under the current rapidly evolving circumstances. Initial reports have suggested that non COVID-19 related cardiovascular mortality and morbidity are likely to be significantly affected [4]. In particular, the number of cardiac surgeries has dramatically decreased as intensive care facilities and staff have been urgently required to treat COVID-19 patients. Even though cardiac surgeons are still required to ensure that essential cardiac interventions are provided to the public, the risk of COVID exposure during hospital admission and its potential impact on surgical outcomes during hospital admission remains uncertain. In healthcare systems where surgeons’ mortality is under public and regulatory bodies scrutiny, such as in the UK, surgeons may be reluctant to offer cardiac operations under the current circumstances. To avoid the risk of inappropriate risk adverse practice, UK regulatory bodies including the SCTS have decided to suspend surgeons’ specific mortality monitoring while national and single unit outcomes remain under strict surveillance.

Anecdotal evidence that patients are reluctant to go to a hospital during the COVID-19 outbreak [3]. Patient’s counselling is particularly challenging as risk stratification methods available do not account for COVID-19 exposure and it takes more time and empathy than ever to help a patient give consent for their cardiac surgery.

In the UK, there are rich resources of routinely clinical data, including the National Adults Cardiac Surgical Audit (NACSA) which will provide essential information on the impact of the COVID-19 pandemic on patients undergoing cardiac surgery. However, clinical observations are accumulating slowly due to drastic reduction of cardiac surgeries performed and data-driven evidence results may not be available until late spring or early fall. As a result, no firm recommendations are still available for case selection and clinical decision making in patients referred to cardiac surgery. In clinical scenarios without compelling evidence, expert consensus can provide information for interim clinical recommendations.

The present survey collected information from the senior cardiac surgeons across all the cardiac units in the UK. First, surgeons agreed that before hospital admission, screening needs to include nasopharyngeal swab, PCR, and chest CT for every patient during the pandemic. Screening is essential to contain the infections and avoid post-operative complications. However, testing before admission can delay treatment which may affect clinical outcomes in some cases and this approach will need to be appraised in the following months. A tailored approach may become necessary if the pandemic continues for several months as suggested by several epidemiologists. Surgeons were also in agreement that the theatre team should adopt full PPE for all the procedures performed during the pandemic. It is possible that the pandemic will last for several months and it will need to be determined the impact of full PPE on team performance (i.e. communication, surgical vision and dexterity and fatigue) and its potential consequences on clinical outcomes. Most surgeons and particularly those working in units currently unaffected by the pandemic, believed that the risk of COVID exposure for patients undergoing cardiac surgery is moderate to high and can have serious consequences on patient’s outcome. This information can represent an important element to support patient’s counselling before compelling clinical evidence becomes available.

Finally, there was a strong consensus that each surgical case requires ad-hoc multidisciplinary team decision and patient’s selection at surgeon’s discretion under the current circumstances was believed to be acceptable only by a very small number of responders. Clearly, multidisciplinary team discussion for each patient requires flexible approaches such as conference call discussions or emails exchanges, and consideration must be given to sensitive data protection and confidentiality and the need of maintaining clinical documentation standards.

There was no strong consensus with regards to specific types of cardiac procedures. However, the majority believed that both aortic and mitral surgery should be considered in selected cases. The role of CABG surgery during the pandemic was more controversial. Neither consensus nor majority was achieved for CABG surgery in selected cases (i.e. left main). Despite recent controversies reported by public media [5], almost a third of responders believing that under the current circumstances percutaneous coronary intervention (PCI) should always be the default strategy for every case and CABG surgery should be considered only in unstable patients when PCI is not feasible. Whether t will have implication on CABG surgery once the pandemic ends remains unclear.

In conclusion, during the COVID-19 pandemic, healthcare policy makers and hospitals not only need to consider methods for containing and treating these infections but how infection outbreaks may affect systems of care beyond the immediate infection. Clinical decision making for patients requiring cardiac surgery is particularly challenging under the COVID-19 pandemic as data-driven evidence is still scarce. Worldwide and in the UK, the lack of firm recommendations for the management of patients requiring cardiac surgery can translate into unwarranted variation in clinical practice and patients’ clinical outcomes across units. In the current unprecedented scenario, systematic appraisal of national and international expert consensus, can represent a rapid and efficient instrument to provide support to heath policy makers and other stakeholders in generating interim recommendations to guide and support clinicians in decision making process.

## Data Availability

Data will be made available on request

## Glossary

AV: aortic valve
CABG: coronary artery bypass graft
COVID-19: Coronavirus disease 2019
CT: computerised tomography
MDT: multidisciplinary team
MV: mitral valve
PRC: polymerase chain reaction
PPEs: Personal Protective Equipment
STEMI: ST-elevation myocardial infarction
TAVI: Transcatheter aortic valve implantation

## Acknowledgement

The authors would like to thank Dr. Arnaldo Dimagli for his invaluable support during the preparation of the present work.

## References

1. Haft JW, Atluri P, Alawadi G, Engelman D, Grant MC, Hassan A, Legare JF, Whitman G, Arora RC; Society of Thoracic Surgeons COVID-19 Taskforce and the Workforce for Adult Cardiac and Vascular Surgery. Adult cardiac surgery during the COVID-19 Pandemic: A Tiered Patient Triage Guidance Statement. Ann Thorac Surg. 2020 Apr 10. pii: S0003-4975(20)30548-8. doi: 10.1016/j.athoracsur.2020.04.003.

2. Hassan A, Arora RC, Adams C, Bouchard D, Cook R, Gunning D, Lamarche Y, Malas T, Moon M, Ouzounian M, Rao V, Rubens F, Tremblay P, Whitlock R, Moss E, Légaré JF; Canadian Society of Cardiac Surgeons. Cardiac surgery in Canada during the COVID-19 Pandemic: A Guidance Statement from the Canadian Society of Cardiac Surgeons. Can J Cardiol. 2020 Apr 8. pii: S0828-282X(20)30323-8. doi: 10.1016/j.cjca.2020.04.001

3. Kea B, Sun BC. Consensus development for healthcare professionals. Intern Emerg Med. 2015;10(3):373–383. doi:10.1007/s11739-014-1156-6

4. Tam CF, Cheung KS, Lam S, Wong A, Yung A, Sze M, Lam YM, Chan C, Tsang TC, Tsui M, Tse HF, Siu CW. Impact of Coronavirus Disease 2019 (COVID-19) Outbreak on ST-Segment-Elevation Myocardial Infarction Care in Hong Kong, China. Circ Cardiovasc Qual Outcomes. 2020 Apr;13(4):e006631. doi: 10.1161/CIRCOUTCOMES.120.006631.

5. https://www.bbc.co.uk/news/health-50715156

6. Ahmad Y, Howard JP, Arnold AD, Cook CM, Prasad M, Ali ZA, Parikh MA, Kosmidou I, Francis DP, Moses JW, Leon MB, Kirtane AJ, Stone GW, Karmpaliotis D. Mortality after drug-eluting stents vs. coronary artery bypass grafting for left main coronary artery disease: a meta-analysis of randomized controlled trials. Eur Heart J. 2020 Mar 2. pii: ehaa135. doi: 10.1093/eurheartj/ehaa135.

